# Neural markers of suppression in impaired binocular vision

**DOI:** 10.1101/2020.09.11.20192047

**Authors:** Freya A. Lygo, Bruno Richard, Alex R. Wade, Antony B. Morland, Daniel H. Baker

## Abstract

**Objective/Purpose:** Even after conventional patching treatment, individuals with a history of amblyopia typically lack good stereo vision. This is often attributed to atypical suppression between the eyes, yet the specific mechanism is still unclear. Guided by computational models of binocular vision, we tested explicit predictions about how neural responses to contrast might differ in individuals with impaired binocular vision.

**Design:** A 5 × 5 factorial repeated measures design was used, in which all participants completed a set of 25 conditions (stimuli of different contrasts shown to the left and right eyes).

**Participants:** 25 individuals with a history of amblyopia, and 19 control participants with typical visual development, participated in the study.

**Methods:** Neural responses to different combinations of contrast in the left and right eyes, were measured using both electroencephalography (EEG) and functional magnetic resonance imaging (fMRI). Stimuli were sinusoidal gratings with a spatial frequency of 3c/deg, flickering at 4Hz. In the fMRI experiment, we also ran population receptive field and retinotopic mapping sequences, and a phase-encoded localiser stimulus, to identify voxels in primary visual cortex (V1) sensitive to the main stimulus.

**Main outcome measures:** The main outcome measures were the signal-to-noise ratio of the steady state visual evoked potential, and the fMRI *β* weights from a general linear model.

**Results:** Neural responses generally increased monotonically with stimulus contrast. When measured with EEG, responses were attenuated in the weaker eye, consistent with a fixed tonic suppression of that eye. When measured with fMRI, a low contrast stimulus in the weaker eye substantially reduced the response to a high contrast stimulus in the stronger eye. This effect was stronger than when the stimulus-eye pairings were reversed, consistent with unbalanced dynamic suppression between the eyes.

**Conclusions:** Measuring neural responses using different methods leads to different conclusions about visual differences in individuals with impaired binocular vision. Both of the atypical suppression effects may relate to binocular perceptual deficits, e.g. in stereopsis, and we anticipate that these measures could be informative for monitoring the progress of treatments aimed at recovering binocular vision.

## Introduction

The binocular visual system is exquisitely sensitive, and has the ability to detect differences (disparities) between the eyes of well below one minute of arc^1^. This results in a vivid perception of depth from stereopsis^2^ that benefits everyday tasks such as fine motor control (e.g. threading a needle) and the judgement of relative object distance (e.g. during driving). But in a substantial minority of individuals (around 1.4%^3^), an optical (e.g. anisometropia) or muscular (e.g. strabismus) asymmetry between the eyes during childhood disrupts the development of binocular vision. This can lead to amblyopia, in which vision through the affected eye is significantly impaired^4^. Such problems can be treated to some extent by orthoptic or surgical interventions, which recover sensitivity in the weaker eye in a proportion of cases^5^. But even if treatment is successful in improving vision in the amblyopic eye, binocular vision may not be restored and stereopsis rarely reaches normal levels^6^. An enduring mystery is the identity of the neural mechanism that disrupts binocular vision, even in situations where the eyes have similar acuity and sensitivity.

In clinical practice, binocular visual disturbances in amblyopia are typically attributed to a process of suppression, whereby the fellow eye suppresses signals from the amblyopic eye^7,8,9^. This suppression could take several different forms. For example, ‘tonic’ suppression should persist even when there is no input to the fellow eye (e.g. if it is closed, patched or pressure blinded, or simply shown a blank display). This amounts to a fixed attenuation of the signal in the amblyopic eye that is invariant to signals from the fellow eye^10^. Alternatively, a more ‘dynamic’ form of suppression would depend on the current stimulation of the two eyes, such that higher contrasts in one eye produce greater suppression of the other eye. Interocular suppression has been widely studied in intact binocular vision, and has several perceptual consequences, such as ocularity invariance (the observation that our general perception of the world is unchanged whether one or both eyes are open^11^) and binocular rivalry (the alternation in perception between conflicting images shown to the two eyes^12^). Impaired binocular vision might result from an imbalance of these existing processes of interocular suppression.

Distinguishing between these, and other, explanations for binocular impairments has proved challenging. In some psychophysical paradigms, such as dichoptic contrast discrimination, similar performance can result even over a wide range of relative amounts of suppression between the eyes^10^. In other paradigms, tonic and dynamic suppression are equally able to account for the results^13^. Isolating a direct neural measure of suppression would allow us to distinguish between different models, and potentially provide an objective index of binocular impairment that could be used to track improvements during treatment. In the present study we measured visual responses to stimuli of different contrasts directly with two methods: functional magnetic resonance imaging (fMRI) and electroencephalography (EEG). These methods have complementary strengths and weaknesses: fMRI has excellent spatial precision, but poor temporal resolution, whereas EEG has poor spatial precision, but good temporal resolution. Previous work measuring visual responses in amblyopia with one or other of these methods has reported generally weaker responses to stimuli in the amblyopic eye^14,15^. However, the two techniques have not previously been directly compared using common stimuli.

Here we use both methods to measure contrast-response functions (Figure 1a) for factorial combinations of contrast shown to the left and right eyes (Figure 2g; see also^11^). We plot the results as a series of functions, where stimuli of increasing contrast are shown to one eye (the ‘target’ stimuli) in the presence of a fixed-contrast (but otherwise identical) stimulus in the other eye (the ‘mask’ stimulus). The general character of these functions can be predicted by contemporary models of binocular signal combination^16,17^, and correspond well to previous measurements in intact binocular visual systems using both fMRI^17^ and EEG^18^. When the mask is absent (0% contrast), the model produces a monotonically increasing monocular contrast response function (left-most curve in Figure 1a). As mask contrast increases, the overall response becomes larger because it combines the target and mask signals together. However, interocular suppression causes a surprising reduction in the response to a high contrast mask when a target of intermediate contrast is added^11,19^. This produces the u-shaped function shown in the final curve of Figure 1a – the response increase caused by excitation is outweighed by the response reduction caused by suppression. Responses therefore go down before they go up, giving a direct measure of interocular suppression.

**Figure 1:**
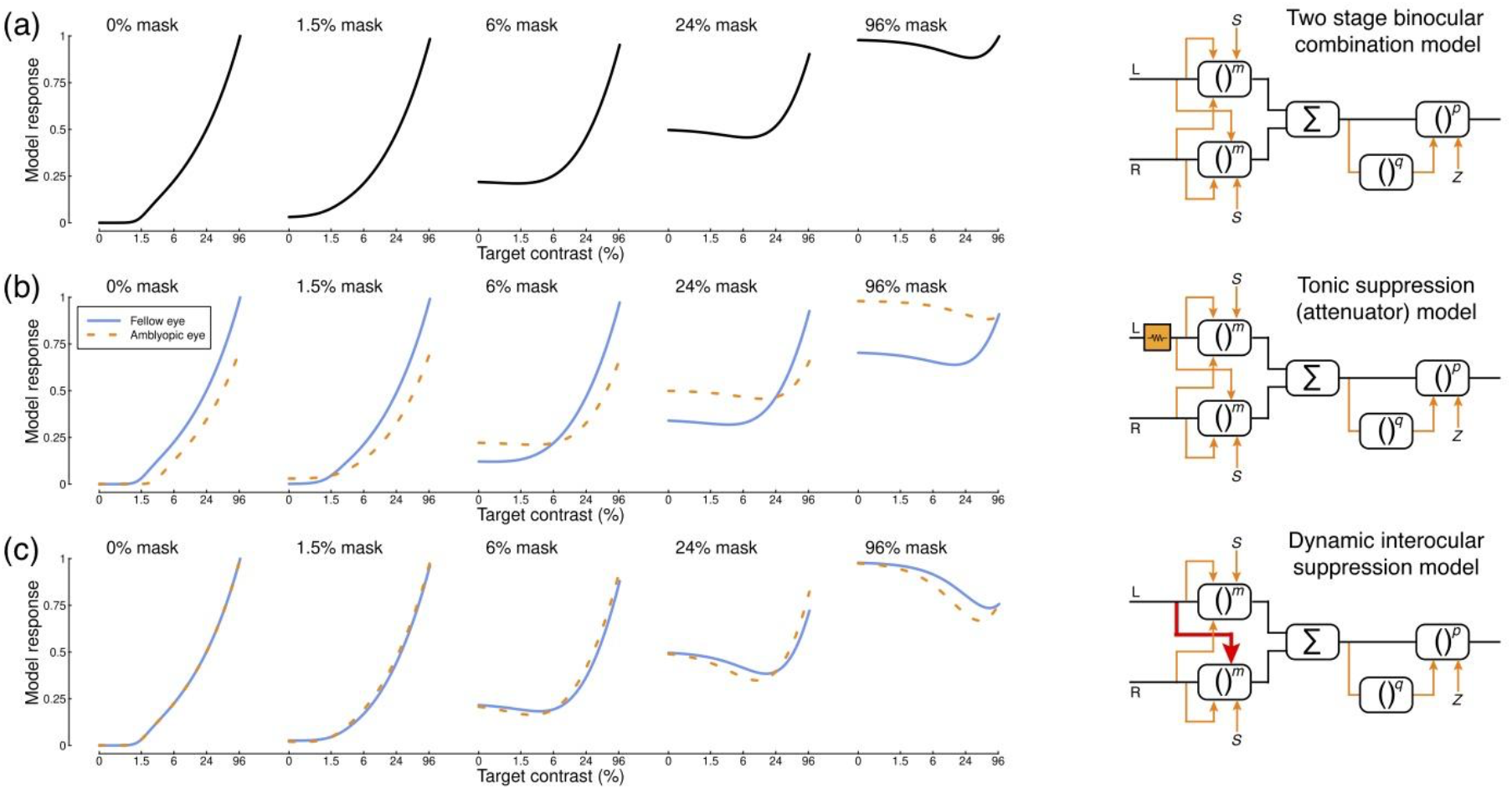
Model predictions and diagrams. Panel (a) shows predictions of the two-stage model (Meese et al., 2006) for different combinations of mask and target contrasts shown to the left and right eyes. The model diagram features multiple stages of gain control (boxes), exponentiation (to powers *m*, *p* and *q)*, inhibition (orange arrows), and binocular summation (denoted by *Σ*). Panel (b) shows a variant of the same model^10^, where the input to the left eye is attenuated prior to any other processing (orange box in the diagram). This affects the model’s behaviour by reducing the response to the affected (e.g. amblyopic) eye (orange dashed curves). Panel (c) shows a further variant in which there is stronger inhibition from one eye onto the other (red arrow in the diagram). This has no effect for monocular stimulation (leftmost function), but increases suppression with high mask contrasts (rightmost functions). Further model details and equations are given in Appendix 1.

**Figure 2:**
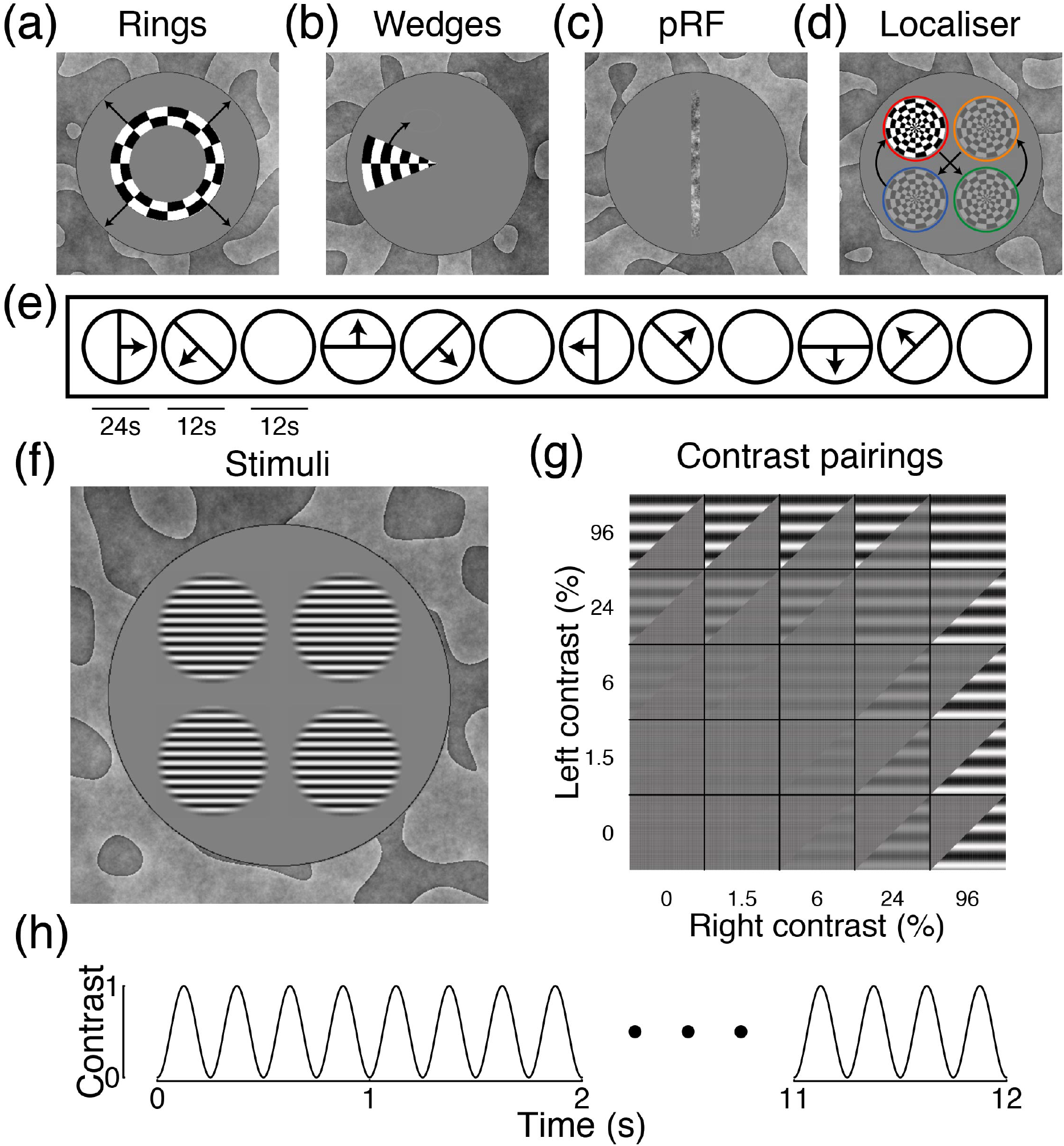
Example stimuli and methodological details. Panels (a,b,c,d,f) show stimuli used in different parts of the study. Panels (a,b) show ring and wedge plaid stimuli used in retinotopic mapping, with black arrows (not presented) indicating the direction of motion. Panel (c) shows the noise bar used in population receptive field (pRF) mapping, which followed the temporal sequence illustrated in panel (e). Panel (d) shows the plaid localiser stimulus, which followed the positional sequence indicated by the arrows (only one plaid was visible in a given 6-second window, and the coloured rings were not shown). Panel (f) shows the sine-wave grating stimuli used to measure contrast response functions. These were presented to the left and right eyes in different contrast combinations, as illustrated in panel (g). The gratings flickered on and off for 12 seconds according to a 4Hz sine-wave, as shown in panel (h).

We can disrupt the model shown in Figure 1a in two key ways. First, we can implement tonic suppression by attenuating the signal in one eye by a constant factor^10^. This reduces the response in the affected eye, and also weakens its impact on the fellow eye (Figure 1b). Second, we can implement dynamic suppression by increasing the weight of suppression from one eye onto the other (Figure 1c). This has no effect on monocular presentations (as there is no signal in the opposite eye to cause suppression), but with high-contrast masks there is a much greater reduction in response for intermediate signal contrasts (the u-shaped functions in the right-most plot become deeper). This experimental paradigm therefore has the potential to distinguish between these two types of suppression.

Our aim in this study was to empirically test specific predictions of these competing models by measuring neural responses with fMRI and EEG to a common set of visual stimuli. In addition to testing control participants with typical binocular vision, we also recruited individuals with a history of binocular disturbance. Although these participants do not all currently meet the diagnostic criteria for amblyopia (owing to successful treatment), they would very likely have done so in childhood and/or had they not been treated. Given the widespread incidence of treatment in countries with developed healthcare systems, understanding the residual binocular deficits in treated amblyopes is of substantial clinical importance. To summarise our results, we find attenuated responses to stimuli in the amblyopic eye when measured using EEG, and increased and asymmetrical interocular suppression in individuals with impaired binocular vision when measured using fMRI. Surprisingly this takes the form of stronger suppression of the dominant eye by the weaker eye.

## Methods

### Participants

A total of 44 adult participants completed the EEG experiment, 19 of whom were control participants with no history of binocular visual abnormalities, and clinically normal vision. The remaining 25 participants had been diagnosed with amblyopia, or treated for strabismus during childhood (see Table 1 for further details). Their ages ranged from 17 to 49, with a mean age of 23.8 years (SD of 7.7 years). Approximately half of these participants (12/25, highlighted in bold) still met the diagnostic criteria for amblyopia at the time of testing, based on a corrected visual acuity difference of two lines or more between the eyes. The MRI experiments were completed by 10 of the control participants, and 12 of the patients (A1 – A12 in Table 1). Participants gave written informed consent, and received financial compensation for their time (£20 per experiment). The study protocols were approved by the research governance committee of the York Neuroimaging Centre, and were consistent with the original wording of the Declaration of Helsinki.

**Table 1:**
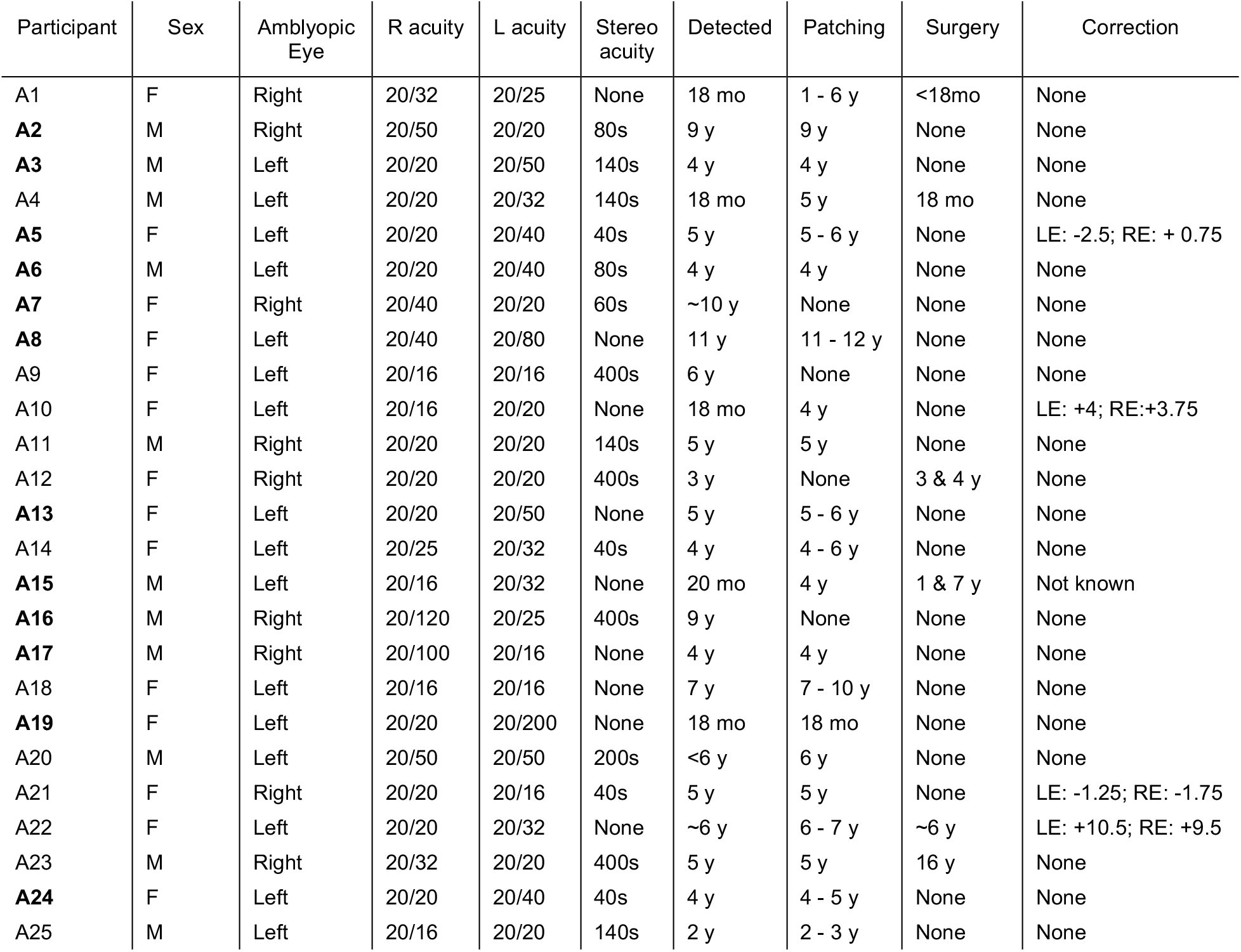
summary of patient demographics, clinical history and acuity measurements. Those highlighted in bold currently meet the clinical criteria for amblyopia (acuity difference of two lines or more on a Snellen chart). None of the participants had a residual strabismus.

### Apparatus and stimuli

In the MRI scanner, stimuli were displayed using a ProPixx DLP projector (VPixx Ltd., Quebec, Canada) with a refresh rate of 120Hz, and a resolution of 1920 × 1080 pixels. Viewed from a distance of 57cm, there were 36 pixels per degree of visual angle. The projector was driven by a high performance PC. A circular polariser interleaved images intended for the left and right eyes (effective refresh rate of 60Hz per eye). Images were projected onto a custom acrylic display panel that maintained the polarisation, and viewed through a front-silvered mirror and passive stereo polarizer glasses. The maximum luminance was 356cd/m^2^ when viewed through the glasses. In the EEG lab, stimuli were displayed using a gamma-corrected ViewPixx 3D LCD display (VPixx Ltd.) with a refresh rate of 120Hz, and a resolution of 1920 × 1080 pixels. Viewed from a distance of 57cm, there were 36 pixels per degree of visual angle. The display was driven by a Mac Pro computer. Active stereo shutter goggles (NVidia 3D Vision), synchronised by an infrared signal, allowed segregation of images to the left and right eyes. Through the goggles, the maximum luminance was 26cd/m^2^. Both display systems had low levels of crosstalk, as measured using a photometer^20^. All experiments were programmed in Matlab, using the Psychophysics Toolbox extensions^21-23^.

Retinotopic mapping involved binocularly presented ring and wedge stimuli constructed from a radial square wave plaid, as illustrated in Figure 2a,b. The plaid had an angular wavelength of 45 degrees (i.e. 8 complete cycles in 360 degrees), a radial frequency of 0.8 cycles per degree, and flickered in counterphase at 4Hz. Expanding rings had a width of 1 plaid cycle, and a period of 12 seconds per sequence (4 ring positions). Rotating wedges were 45 degrees (1 cycle) wide, with a period of 24 seconds per rotation (8 positions in 45 degree clockwise steps). Population receptive field (pRF) mapping used a drifting bar (0.5 × 10 degrees) of dynamic 1/f noise, with an RMS contrast of 0.2 (see Figure 2c). The bar drifted at a speed of 0.4 deg/sec, and followed the sequence illustrated in Figure 2e. The pRF stimulus was presented to either the left or right eye in different blocks, with the other eye viewing mean luminance. We used a phase-encoded localiser stimulus, constructed from a radial plaid with a width of 4 degrees (see Figure 2d), presented binocularly. The localiser stimulus counterphase flickered at 4Hz, and changed position every 6 seconds, according to the sequence illustrated in Figure 2d. Each stimulus location had an x-y offset of ±2.34 degrees from fixation. Stimuli for the main experiments were four horizontal sine-wave gratings with a spatial frequency of 3c/deg, a cosine-blurred spatial window, and a width of 4 degrees (see Figure 2f). Five different Michelson contrast values (defined as 100*(L_max_-L_min_)/(L_max_+L_min_), where L is luminance) were presented in different combinations (see Figure 2g). The stimuli flickered sinusoidally between 0 and their nominal contrast (on/off flicker) at a frequency of 4Hz (see Figure 2h). The grating stimuli had x-y offsets of ±2.34 degrees from fixation. In all experiments, a static binocular texture was presented to aid fusion. This was constructed from low spatial frequency bandpass filtered noise, and filled the display beyond the central 12 degree stimulus aperture (see Figure 2 for examples).

### MRI acquisition

All MRI data were acquired using a GE 3T HDx Excite MRI scanner. We collected two high resolution T1-weighted structural scans (TR 7.8 ms; TE 3 ms; voxel size 1 × 1 × 1 mm; 12° flip angle; matrix size 256 × 256; FOV 256 mm), and two T2*-weighted fast gradient recalled echo scans (TR 400 ms; TE 4.2 ms; voxel size 1 × 1 × 2 mm; 25° flip angle; matrix size 128 × 128; FOV 260 × 260 mm), using an 8-channel surface coil (Nova Medical, WiL_min_gton, MA, USA). We acquired functional images using an EPI sequence with a 16-channel posterior surface coil (Nova Medical, WiLmington, MA, USA), to optimize signal-to-noise ratio at the occipital pole. The slice prescription covered the region containing the calcarine sulcus and occipital pole with 39 axial slices (TR 3000 ms; TE 30 ms; voxel size 2 × 2 × 2 mm; 90° flip angle; matrix size 96 × 96; FOV 192 × 192 mm). We also acquired an in-plane proton density scan using the same slice prescription to aid alignment with the structural scans.

Participants completed the MRI experiments in two sessions. In the first session, we collected the structural scans, and the retinotopic mapping and pRF data. Each pRF sequence lasted 396 seconds (132 TRs), and either the left or right eye was stimulated. Two repetitions for each eye were completed. The retinotopic mapping (ring and wedge) sequences were collected as a single scan lasting 204 seconds (68 TRs). In the second session, the phase-encoded localiser and contrast response function data were collected. The localiser scan lasted 156 seconds (52 TRs), and consisted of a blank period (12 seconds), followed by 6 repetitions of the localiser sequence. The contrast response function sequence lasted 612 seconds (204 TRs), and tested each of the 25 conditions (see Figure 2g) once, with 12-second blank periods between each 12-second trial. This was repeated four times for each participant. During all functional scans, participants performed a fixation task, in which they monitored a grid of 9 squares (3×3, each 0.14 degrees wide) with random luminances in the centre of the screen. They were instructed to press a button whenever the fixation marker was changed by re-randomising the luminances. This occurred at randomly determined times, on average once every 48 seconds (i.e. once every two trials). The task was intended to maintain attention and fixation, and we did not record the responses.

### MRI analysis

Primary MRI analysis was conducted in Matlab using the mrVista toolbox (https://github.com/vistalab/vistasoft). Functional data were motion-corrected within and between scans, and aligned to the in-plane (proton density) scan, and subsequently to the participant’s anatomical space. The first 12 seconds (4 TRs) of each functional scan were discarded to account for magnetic saturation effects. Structural scans were processed using Freesurfer^24,25^ to generate a 3D model of the cortex. We created flat patches of unfolded cortex for each hemisphere (120 mm in diameter), centred on the occipital pole (see Figure 3) to facilitate data visualisation and the creation of Regions of Interest (ROIs). The ring and wedge retinotopy and localiser scans were summarised by a coherence (travelling wave) analysis^26^ to calculate the phase of the BOLD response at the repetition frequency of the stimulus for each voxel. The pRF data were fit by estimating (at each voxel) the parameters of a 2D Gaussian function that best predicted the BOLD timecourse, given the position of the bar stimulus^27^. This was done independently for the left and right eye scans. The contrast response function data were combined across repetition and analysed using a general linear model (GLM), with regressors (8 weights) for each of the 25 conditions. We used a combination of retinotopy and pRF results to define a V1 ROI on the flattened cortex using the location of the calcarine sulcus and reversals of phase angle. ROIs were further restricted using the localiser data, by retaining voxels with a coherence exceeding 0.3. Results were saved as Matlab files, and imported into *R* for statistical analysis and visualisation. We also converted GLM *β* weights to MNI space (using tools from FSL^28^), and averaged them across participants for visualisation on an inflated cortex in the Connectome Workbench software^29^.

**Figure 3:**
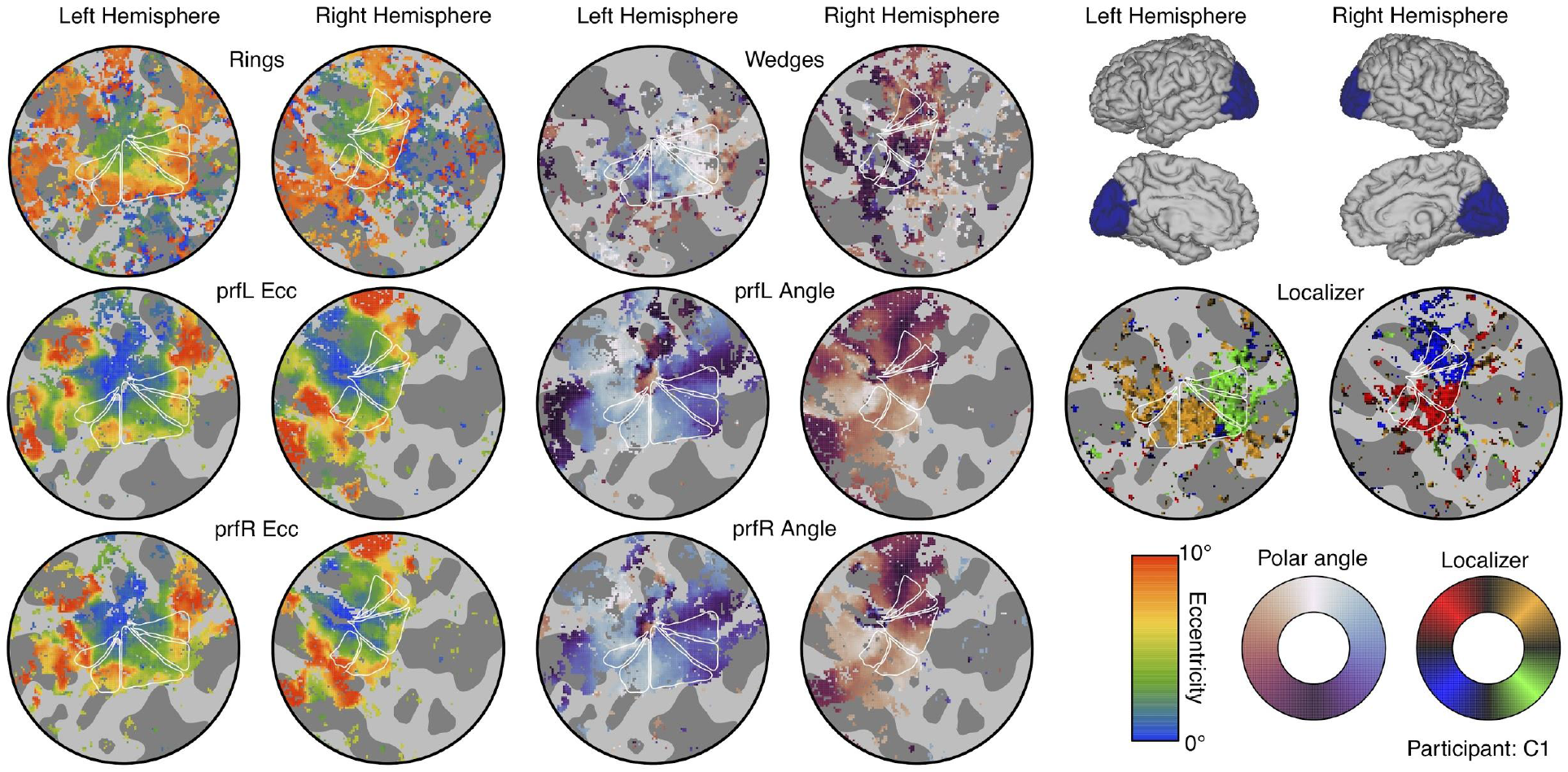
Summary of retinotopic mapping and functional localizer results. The icons in the upper right corner indicate the region of cortex in each hemisphere used to make the flat maps (each 120 mm in diameter). The left-most columns show eccentricity estimates from a plaid ring localiser (top row), and pRF models for stimuli shown to the left (middle row) and right (lower row) eyes. The middle columns show the polar angle parameter from a rotating plaid wedge localiser (top row) and the two pRF models (middle and lower rows). The flat maps in the right-most columns show the responses to the phase-encoded plaid localiser. Colour maps for each measure are shown in the lower right corner. In each flat map, dark grey regions indicate sulci, and white triangles show the locations of visual areas V1 (middle triangle) and V2/3v/d (outer triangles). The phase encoded retinotopy and localiser results were thresholded at a coherence value of 0.3, and the pRF results were thresholded at 10% of explained variance.

### EEG acquisition

All EEG data were acquired using a 64-channel ANT Neuroscan system, with electrodes positioned in a Waveguard cap according to the 10-20 system. Signals were recorded at 1000Hz, and referenced to the whole-head average. Low-latency digital triggers were sent from the stimulus computer to the EEG amplifier using a parallel cable, and recorded stimulus onset and condition codes to the EEG trace. Participants completed 8 repetitions of the contrast response function experiment. On each repetition, stimuli were presented for trials of 12 seconds, with an intertrial interval of 3 seconds. All 25 conditions (see Figure 2g) were presented once per repetition in a random order, taking 375 seconds per block. Participants were given breaks between blocks. The same fixation task as described for the MRI experiments was performed throughout the experiment to maintain attention.

### EEG analysis

Raw data were converted to a compressed csv format using functions from EEGlab^30^, and were then imported into *R* for analysis. We took the Fourier transform of the EEG waveform at each electrode, for a ten-second window beginning one second after stimulus onset (to avoid onset transients). Fourier spectra were averaged across four occipital electrodes *(Oz, POz, O1* and *O2*), and across repetition, using coherent averaging (i.e. retaining the phase information). We then calculated signal-to-noise ratios (SNRs) by dividing the absolute amplitude in the signal bin (4Hz) by the mean of the ten adjacent bins (±0.5Hz in steps of 0.1Hz). These SNRs were averaged across participants, and standard errors were calculated using bootstrapping. To plot the timecourse of SSVEP activity, we repeated the Fourier transform using a sliding one-second window (in steps of 10ms), and scaled by the two adjacent bins (±1 Hz) to calculate the SNR.

### Data and script availability

Data, experiment code (in Matlab) and analysis code (in Matlab and R) is available at: http://dx.doi.org/10.17605/OSF.IO/X9ZR8

## Results

We used the results from retinotopic mapping scans with ring and wedge stimuli (Figure 2a,b), and population receptive field (pRF) sequences (Figure 2c,e), to identify primary visual cortex (V1) on flattened discs of occipital cortex for each hemisphere. Example flat maps are shown in Figure 3 for one control participant (see the project repository for equivalent plots for all participants: https://osf.io/x9zr8/). The phase angle of the BOLD response to the ring stimuli, and the pRF eccentricity values, showed a typical central-to-peripheral gradient^27^, and were highly consistent (e.g. across left and right eye pRF sequences). The phase angle across the wedge and pRF scans showed strong hemisphere/hemi-field segregation (i.e. the right hemisphere responded to stimuli in the left hemi-field and vice versa), as well as the expected phase reversals^31^ that were used to determine boundaries between V1, V2 and V3 (shown by the white triangles in each map – the middle triangle is V1). The V1 region-of-interest (ROI) was further restricted using the responses to a phase-encoded localiser stimulus (see Figure 2d). These responses also showed strong hemisphere/hemi-field and dorsal/ventral segregation, and we retained voxels in the V1 ROI that produced responses with a coherence of 0.3 or higher (right-most flat maps in Figure 3). This resulted in a mean total V1 ROI size (combined across hemispheres) of 601 voxels, and no significant difference in ROI size between patients and controls (*t*(20)=0.54, *p*=0.60).

The localiser-restricted V1 ROIs for each participant were then used to estimate neural responses to stimuli of different contrasts presented to the two eyes (Figure 2f). The BOLD response in this ROI had a typical timecourse^32^, which was modulated by stimulus contrast (see Figure 4a). We fitted a general linear model (GLM) to the full timecourse to estimate a *β* coefficient for each stimulus condition at each voxel. The *β* weights were strongly modulated by stimulus contrast at the occipital pole (red shading in Figure 4b). In a separate experiment using identical stimuli, we also recorded steady-state visual evoked potentials (SSVEPs) using EEG. These showed clear modulation of signal-to-noise ratio (SNR) with stimulus contrast at the flicker frequency of the stimuli (4Hz, Figure 2g), but with a less sluggish timecourse than the BOLD response (see Figure 4c). The responses were well-isolated in the Fourier amplitude spectrum (see Figure 4d), and localised to occipital electrodes (Figure 4d inset). We therefore used the fMRI *β* weights averaged across the ROI, and the SSVEP SNRs averaged across four occipital electrodes (black points in the Figure 4d inset) to calculate contrast response functions for each experiment. Exploratory analyses in V2 and V3, and using different localiser thresholds to define the V1 ROI, produced very similar functions (not shown), as did using raw amplitude values for the EEG experiment (rather than SNR values).

**Figure 4:**
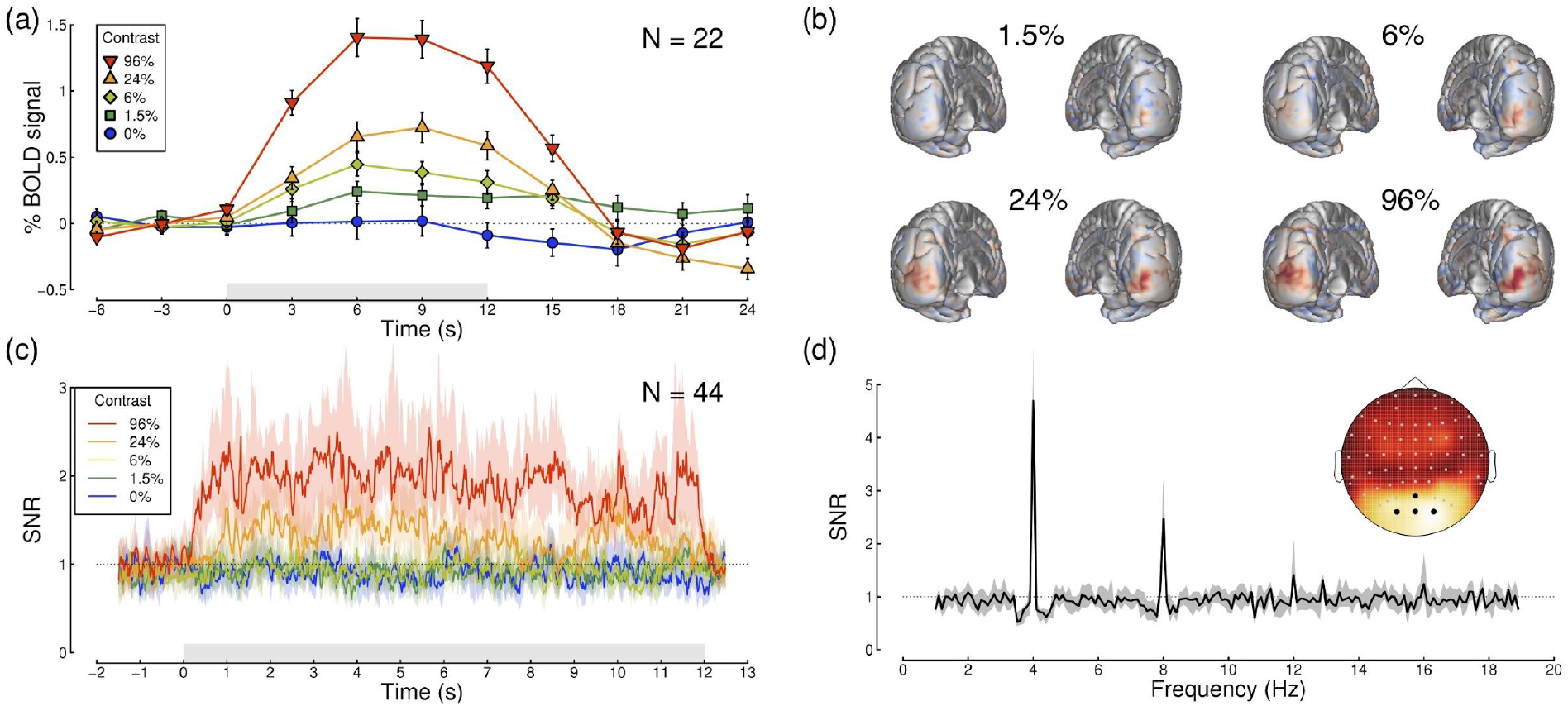
Timecourses and topographies of visual responses measured using fMRI and EEG. Panel (a) shows the BOLD timecourse in the localiser-restricted V1 ROI, for binocular presentation at five stimulus contrasts (see legend). The grey rectangle adjacent to the x-axis indicates the period when the stimulus was presented, and error bars indicate ±1SE across participants (N=22). Panel (b) shows averaged beta weights (unthresholded) from the general linear model, projected on a posterior view of each hemisphere, for the non-zero stimulus contrasts averaged across all participants (subtracting the 0% condition as a baseline). Panel (c) shows the SSVEP timecourse as a signal-to-noise ratio (SNR) at the target flicker frequency (4Hz), calculated using a 1000ms sliding window (centred at the time indicated on the x-axis). Shaded regions indicate bootstrapped 95% confidence intervals of the median, calculated across participants (N=44). The grey shaded rectangle represents the period when the stimulus was presented. Panel (d) shows the Fourier spectrum for a 96% contrast binocular target, averaged across all participants (N=44), for 10-second windows of stimulation. Clear peaks in SNR are apparent at the stimulus flicker frequency (4Hz) and its harmonics (especially 8Hz). The grey shaded region indicates 95% confidence intervals of the median. The inset scalp topography shows that responses were strongest at posterior electrode sites over early visual areas. For panels (c,d), signals were averaged across the electrodes indicated in black on the scalp plot (*Oz*, *POz, O1* and *O2)*.

Figure 5 shows contrast response functions from both experiments, split by participant group (control participants in Figure 5a,c, patients in Figure 5b,d). The first contrast response function in each row (circle symbols) is for monocular stimulus presentation, and shows the expected monotonic increase for each data set. As predicted by our computational models (see Figure 1), as mask contrast increased, the functions rose from baseline across the plot. Statistically, there were significant main effects of both target and mask contrast, and significant interactions, for all data sets (see Table 2). For the patients there was a very slight reduction of response in the amblyopic eye (orange circles) compared with the fellow eye (blue circles) at the highest target contrast in the fMRI data (Figure 5b), though this was not significant (*t*(11)=1.20, *p*=0.26, *d*=0.35). The difference was more pronounced in the SSVEP data (Figure 5d), and was significant at 24% target contrast (*t*(24)=2.91, *p*<0.01, *d*=0.58), though not at 96% contrast (*t*(24)=1.96, *p*=0.06, *d*=0.39).

**Figure 5:**
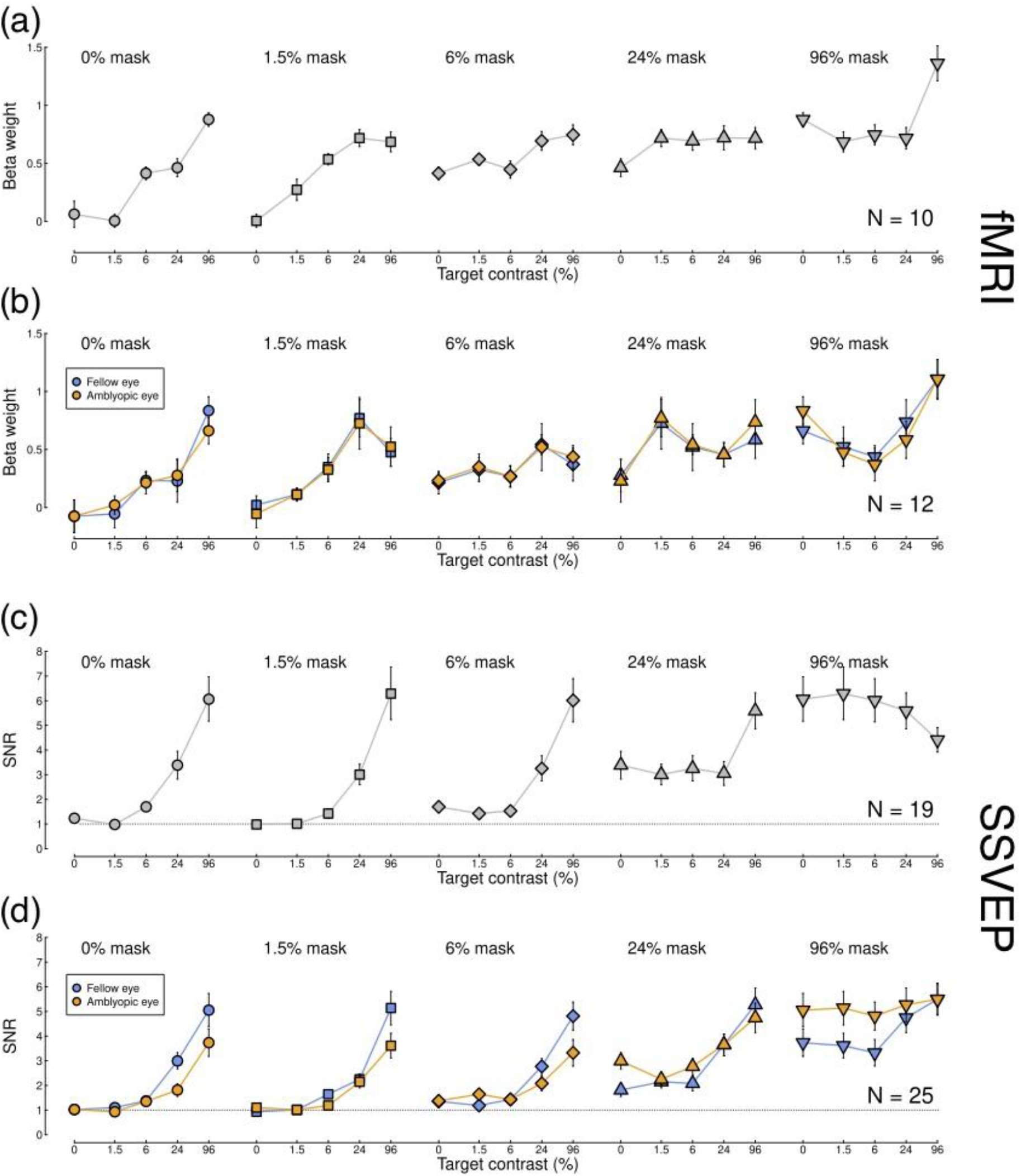
Contrast response functions measured using fMRI and EEG. Data for control participants (panels a,c) are averaged across complementary conditions for the left and right eyes. Data for patients (panels b,d) are plotted considering the fellow eye as the ‘target’ eye (blue) and also considering the amblyopic eye as the ‘target’ eye (orange). These data are identical, but are re-ordered to aid interpretation. Error bars in each plot indicate ±1SE across participants.

**Table 2:** ANOVA results for contrast response functions.

| Data set | Effect | F-ratio (df) | p-value | Effect size ($\omega^2$) |
| --- | --- | --- | --- | --- |
| Control fMRI | Left eye contrast | 18.82 (4,36) | <0.001 | 0.27 |
| Control fMRI | Right eye contrast | 19.56 (4,36) | <0.001 | 0.22 |
| Control fMRI | Interaction | 2.91 (16,144) | <0.001 | 0.18 |
| Patient fMRI | Fellow eye contrast | 12.80 (4,44) | <0.001 | 0.14 |
| Patient fMRI | Amblyopic eye contrast | 8.47 (4,44) | <0.001 | 0.14 |
| Patient fMRI | Interaction | 2.92 (16,176) | <0.001 | 0.15 |
| Control SSVEP | Left eye contrast | 29.93 (4,72) | <0.001 | 0.31 |
| Control SSVEP | Right eye contrast | 27.21 (4,72) | <0.001 | 0.28 |
| Control SSVEP | Interaction | 6.43 (16,288) | <0.001 | 0.14 |
| Patient SSVEP | Fellow eye contrast | 34.88 (4,96) | <0.001 | 0.38 |
| Patient SSVEP | Amblyopic eye contrast | 19.26 (4,96) | <0.001 | 0.17 |
| Patient SSVEP | Interaction | 2.96 (16,384) | <0.001 | 0.04 |

For the control participants, there was evidence of interocular suppression (a u-shaped function) for the highest mask contrast when measured using fMRI (final function in Figure 5a), though this was not statistically significant (paired samples t-test comparing 0% and 6% target contrast conditions when a 96% contrast mask was present, *t*(9)=1.8 *p*=0.10, *d*=0.57, mean difference of 0.13 *β* units). For the control EEG data interocular suppression occurred at a higher target contrast (96%, see final function in Figure 5c), but was also not significant (*t*(18)=0.96, *p*=0.35, *d*=0.22, mean difference of 0.44 SNR units). The u-shaped function was more pronounced in the patients (Figure 5b,d). For the fMRI data, suppression was more substantial when the mask was shown to the fellow eye (orange inverted triangles comparing 0% and 6% target levels with a 96% mask; *t*(11)=3.25, *p*=0.008, *d*=0.94, mean difference of 0.46 *β* units) than the amblyopic eye (blue inverted triangles; *t*(11)=2.51, *p*=0.029, *d*=0.72, mean difference of 0.22 *β* units). A very subtle suppression effect was qualitatively apparent for the SSVEP data (Figure 5d), but this did not reach statistical significance for either eye (both p>0.05). SSVEP responses at the second harmonic frequency (8Hz) were broadly similar to those at the fundamental (see Supplementary Figure S1).

To further investigate the interocular suppression effect, we plotted full BOLD timecourses for the condition where the mask only was shown (96% contrast mask to one eye, 0% contrast target to the other), and the condition where the same mask was paired with a 6% contrast target in the other eye (see Figure 6). Interocular suppression is clear in each data set, as the white points appear below the black points over much of the function. Our computational model (see right-most functions in Figure 1a,c) predicts that this happens because the excitatory impact of the 6% contrast stimulus is outweighed by its suppression of the response to the 96% contrast stimulus in the other eye: overall activity goes down instead of up. However this effect is much more substantial for the fellow eye of the patients (Figure 6b) than for the amblyopic eye (Figure 6c), consistent with our finding from the *β* weights (Figure 5b). The cortical meshes along the upper row of Figure 6 show the difference in *β* weights between conditions, with blue shading indicating stronger suppression. Suppression is apparent at the occipital pole, and is again strongest for the fellow eye of the patients (Figure 6b).

**Figure 6:**
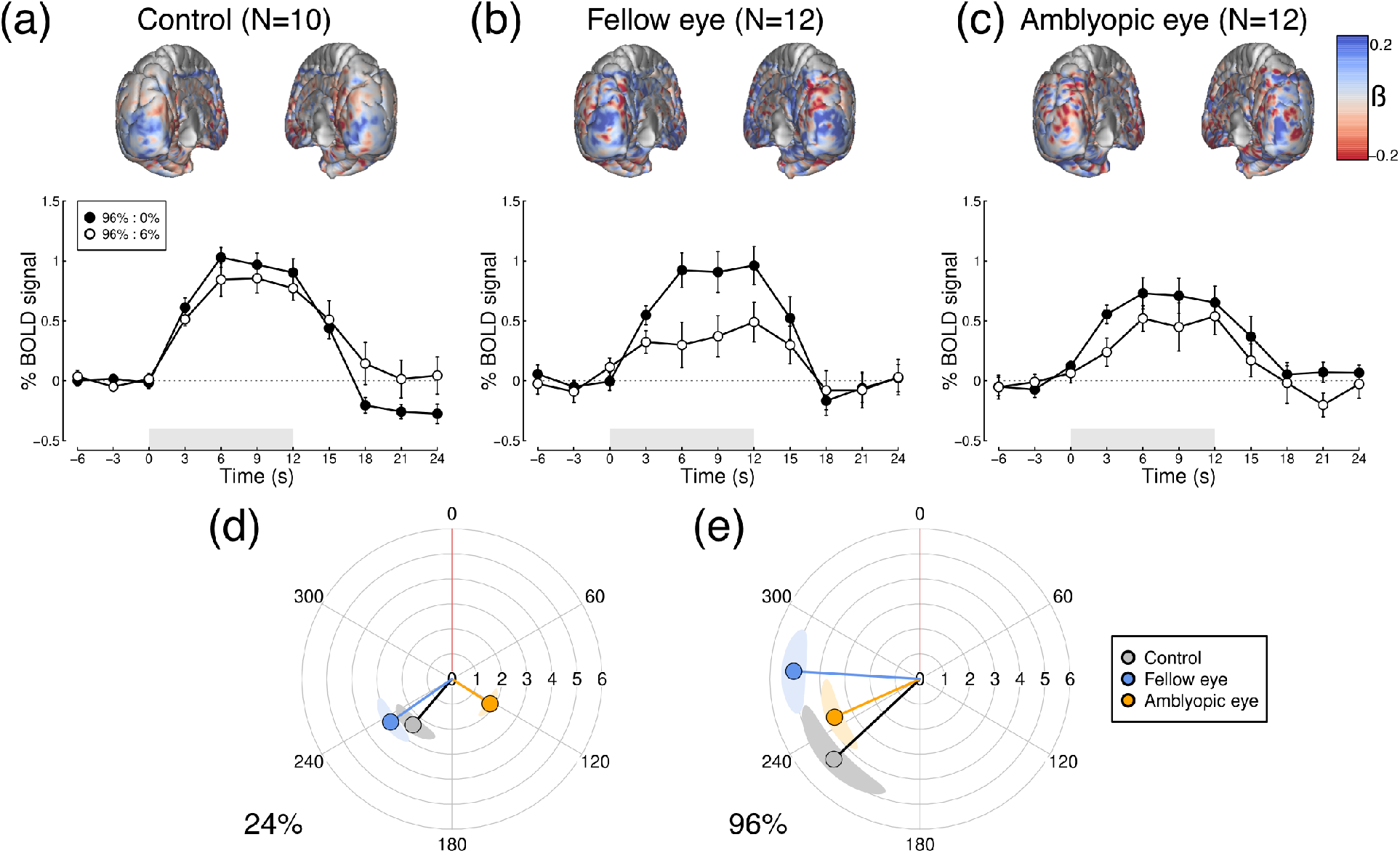
Timecourse and spatial distribution of interocular suppression measured using fMRI, and polar plots of SSVEP responses to monocular stimuli. Upper plots show the difference of *β* weights at each voxel (unthresholded), between a monocular condition where one eye saw 96% contrast, and a dichoptic condition where the eyes saw 96% and 6% contrast. Blue shading indicates a suppressive effect (and red shading a facilitatory effect) of the 6% component. Lower panels show the timecourse of % signal change for the same two conditions. Data are shown for (a) control participants, (b) the 96% contrast mask in the fellow eye of the patients, and (c) the 96% contrast mask in the amblyopic eye of the patients. Error bars indicate ±1SE across participants, and grey shaded rectangles show the duration of stimulus presentation. Panels (d,e) show SSVEP responses for monocularly presented stimuli at 24% (d) and 96% (e) contrast. This representation shows a phase lag (i.e. angular difference) between the amblyopic (orange) and fellow (blue) eyes. Shaded regions indicate ±1SE, calculated independently for amplitude and phase values.

The lower two panels of Figure 6 show polar plots comparing monocular SSVEP responses to stimuli of 24% contrast (Fig 6d), and 96% contrast (Fig 6e). There is a phase lag between the fellow and amblyopic eye in both panels. For the 24% contrast target, this is approximately 112 degrees, which corresponds to a lag of around 78 ms at the 4Hz flicker frequency used here. For the 96% contrast, the lag is around 28 degrees (20 ms). This latter estimate corresponds well with previously reported phase lags using a similar paradigm in magnetoencephalography (MEG)^33^.

## Discussion

We measured neural responses to different combinations of contrast in the left and right eyes, using both EEG and fMRI. In participants with atypical binocular vision, we found reduced responses in the amblyopic eye using EEG, and increased suppression between the eyes (compared with controls) in V1 using fMRI. These different effects are consistent with greater tonic and dynamic suppression (respectively) in individuals with impaired binocular vision, and may be responsible for the deficits in stereopsis experienced by the majority of these participants (see Table 1). We now discuss why the results differ across measurement methods, what these findings tell us about amblyopic suppression, and how treatments might be targeted towards the development of functioning binocular vision.

### Comparison of EEG and fMRI measures

This is the first study to use both EEG and fMRI to investigate contrast processing in impaired binocular vision. Previous studies using either EEG or MEG^14,33^ have typically found larger amblyopic deficits in the monocular response than those using fMRI^34,35^ (though some work has shown substantial fMRI deficits^36^), mirroring our results here (left-most functions of Figure 5b,d). Considering only this previous work, heterogeneity of stimuli and participants across studies might well have explained the differences. However in the present experiments we used identical stimuli, and the same participants completed both experiments (Supplementary Figure S2 shows the EEG data for only the participants who completed the MRI experiments). The reduction in response amplitude to stimuli in the weaker eye is clearer using SSVEP than using fMRI. However, differences in dynamic interocular suppression are only apparent when measured with fMRI (right-most functions in Figure 5a,b; Figure 6a-c).

Analogous differences between these two methods have recently been reported in the study of attention. Itthipuripat et al.^37^ found that spatial attention produces a change in baseline response when measured using fMRI, but not when measured using EEG (including evoked potentials and SSVEPs). Instead fMRI, which measures global neural activity indirectly via oxygen consumption, appears to be more closely related to late ERP components and alpha band oscillations recorded using EEG^37^, rather than the stimulus onset transients detected by SSVEP and early ERP components. Although our contrast response functions do not involve baseline shifts, the other differences between our fMRI and EEG results are consistent with different features of neural activity being probed by these two methods.

One possible explanation for the differences in monocular response between methods is that the firing of visual neurons responsive to the amblyopic eye might be desynchronised. This would have a greater effect on phase-locked SSVEP responses – which depend upon synchronised firing – than on fMRI BOLD responses, which are a proxy for overall neural activity. Instead, asynchronous activity in higher frequency bands (50 – 200Hz) shows a closer correspondence with the BOLD response^38^, and these signals can be detected with extracranial techniques such as MEG^39^. In terms of the differences in suppression, this could reflect processing in different layers of cortex. Evoked responses measured using EEG correspond most closely to activity in the more superficial (supragranular) layers of cortex, whereas much inhibitory processing involves deeper (granular) layers^40^. Future work using laminar fMRI at higher field strength (7T or above^41^), or multi-unit electrophysiology^42,43^, could allow dissociation of excitatory and inhibitory responses in amblyopia. In addition, these methods can resolve ocular dominance columns in V1, allowing eye-specific inputs to be measured directly.

### What is amblyopic suppression?

Suppression in amblyopia is measured very differently in clinical practice compared with the methods used in lab-based neuroscience research. The most common clinical measures are the Worth 4-dot test, and Bagolini striated lenses, both of which give a qualitative indication of whether one eye’s image is substantively suppressed by the other eye during binocular viewing. Lab-based measures involve a variety of paradigms, including psychophysical approaches such as dichoptic masking^10,44-46^, or assessing binocular fusion of edges^47^ or gratings^48^, and more direct neurophysiological estimates of suppression in animal models^49,50^. However, because both eyes are being stimulated during testing, clinical suppression could in principle be explained by either tonic or dynamic suppression, and most psychophysical work also cannot distinguish between these possibilities. Many of our patients (around half) no longer met the clinical criterion for amblyopia, yet as a group they still exhibited greater dynamic interocular suppression than our control participants, as well as tonic suppression of one eye. The finding that both types of suppression are apparent, even in individuals who have received patching or surgical interventions, strongly suggests that both will also be present in untreated amblyopes. Amblyopic suppression might therefore involve a combination of these two processes.

One feature that remains to be determined is whether one type of suppression is a primary cause of amblyopia, and the other a later consequence. Cortical suppression, characterised as a process of gain control^51^, is a dynamic, adaptive process that acts to optimise the sensory response^52^. It is clear that binocular vision is particularly plastic, as the relative weighting of the two eyes can be altered following a brief period of occlusion^53^, and this may be the mechanism by which clinical patching treatment improves vision. Our finding of increased suppression of the fellow eye by the amblyopic eye could be an attempt to rebalance an asymmetrical system. If so, this might influence the development of novel treatments geared towards modulating interocular suppression.

### Perceptual consequences of suppression, and potential for targeted treatment

It is generally assumed that clinical suppression results in viewing the world ‘through’ the fellow eye, with signals from the amblyopic eye being completely suppressed. However, other work has shown that when signals in the amblyopic eye are boosted by an appropriate amount, information is still summed binocularly^54^. Indeed, the principle of ocularity invariance (that the world does not change when one eye is closed) makes it very difficult to distinguish between suppression and fusion outside of the laboratory or clinic. However, even if the amblyopic eye still contributes to perception, stereopsis is extremely sensitive to imbalances between the eyes, and breaks down when one eye receives a stronger input^55^. The two types of suppression we identify here would likely unbalance signals in exactly this way, which may contribute to the poorer stereopsis for most of our patients (see Table 1).

Consistent with this idea, several treatments have recently been proposed that aim to reduce suppression between the eyes. For example, antisuppression therapy^56^ involves presenting dot motion stimuli, in which signals are shown to one eye, and noise distractors to the other. To perform the task, patients must favour information from the signal (amblyopic) eye. Performance improves over time, implying that suppression is reduced. This approach also improves acuity and stereopsis, even in amblyopic adults far beyond the critical period (in childhood) for traditional treatment. Related treatments that involve playing dichoptic video games^57-59^, or watching dichoptic movies^60^, may work in a similar way. Measuring the two types of suppression identified here throughout treatment would reveal causal relationships between suppression and visual function, potentially allowing clinicians to optimise treatment schedules and monitor progress.

## Data Availability

Data, experiment code (in Matlab) and analysis code (in Matlab and R) is available at:
http://dx.doi.org/10.17605/OSF.IO/X9ZR8

## Appendix 1 details of computational models

The two stage model of Meese et al.^16^ comprises an intial stage of monocular gain control, followed by binocular summation, defined as:

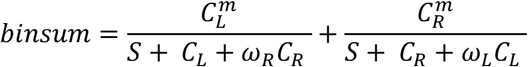

where *m* = 1.3, *S* = 1, and *ω_R_ = ω_L_* = 1, *C_L_* and *C_R_* are the input contrasts to the left and right eyes. The output of the model follows a further nonlinearity:

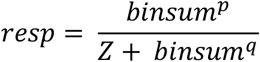

where *p* = 8, *q* = 6.5, and *Z* = 0.1. The predictions shown in Figure 1a are the output of the model for different combinations of *C_L_* and *C_R_*. To predict the effects of attenuating one eye’s input (Figure 1b), an attenuator parameter^10^ is added to one eye’s input:

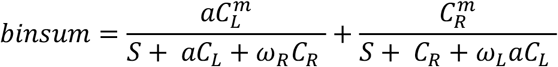

where *a* = 0.5. Finally, to assess the impact of unbalanced interocular suppression (Figure 1c), we set *ω_R_* = 2, but left *ω_L_* = 1 (and set *a* = 1). Precise parameter values, e.g. of exponents, are not essential to produce the overall model behaviour.

## Precis

Evidence of atypical interocular suppression in individuals with impaired binocular vision was obtained using EEG and fMRI, with a single experimental paradigm. This may underlie the deficits in stereopsis experienced by treated amblyopes.

## Acknowledgements

We are grateful to all of our participants for their involvement in this work, and to Robert Hess for helpful discussions on suppression in amblyopia.

## Online Supplemental materials

**Figure S1:**
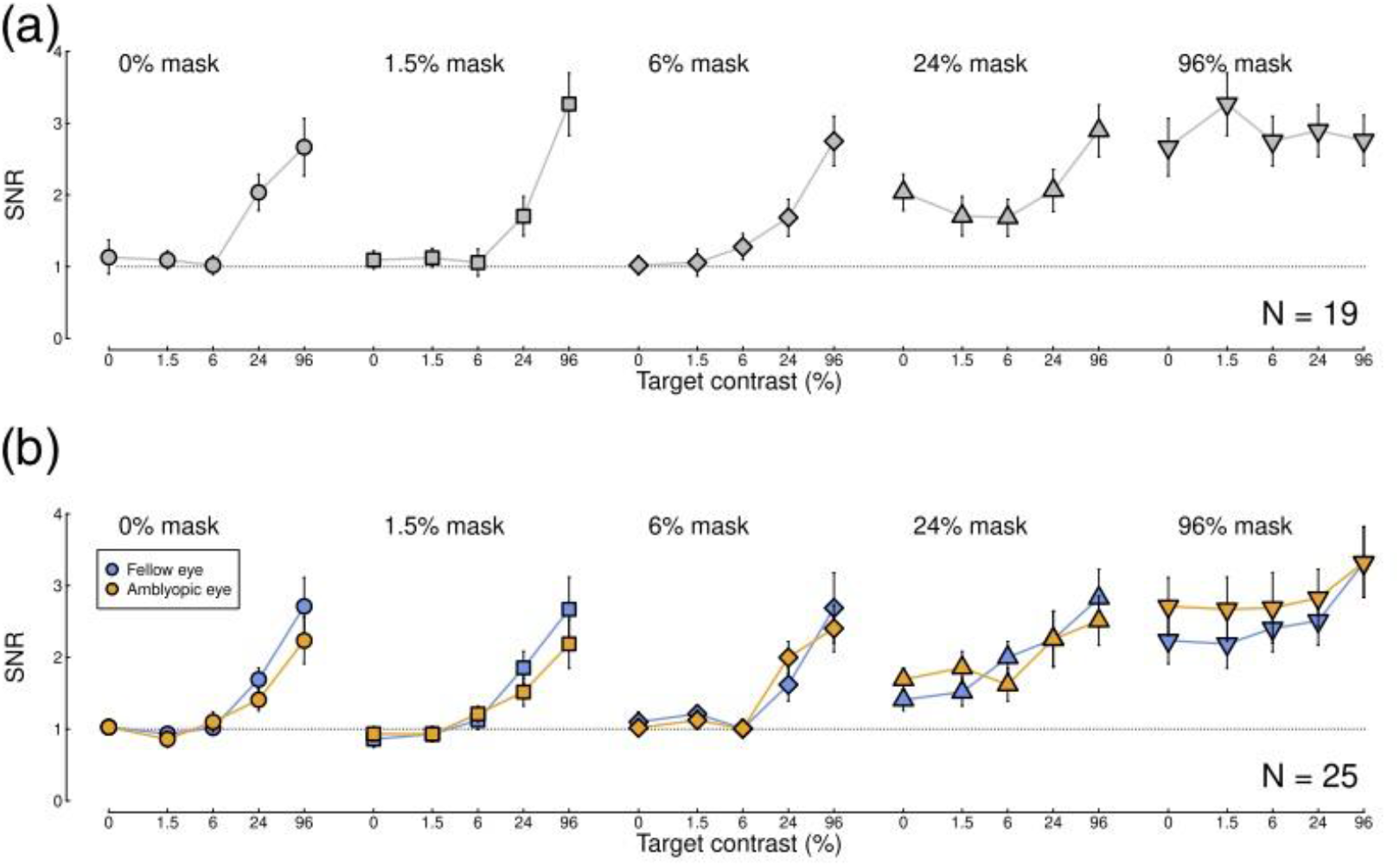
SSVEP data for the second harmonic (2F) response. The main trends are the same as in Figure 5c,d, though the SNRs are overall lower (note y-axis scaling). Panel (a) shows data from control participants (N=19), and panel (b) shows data from patients (N=25).

**Figure S2:**
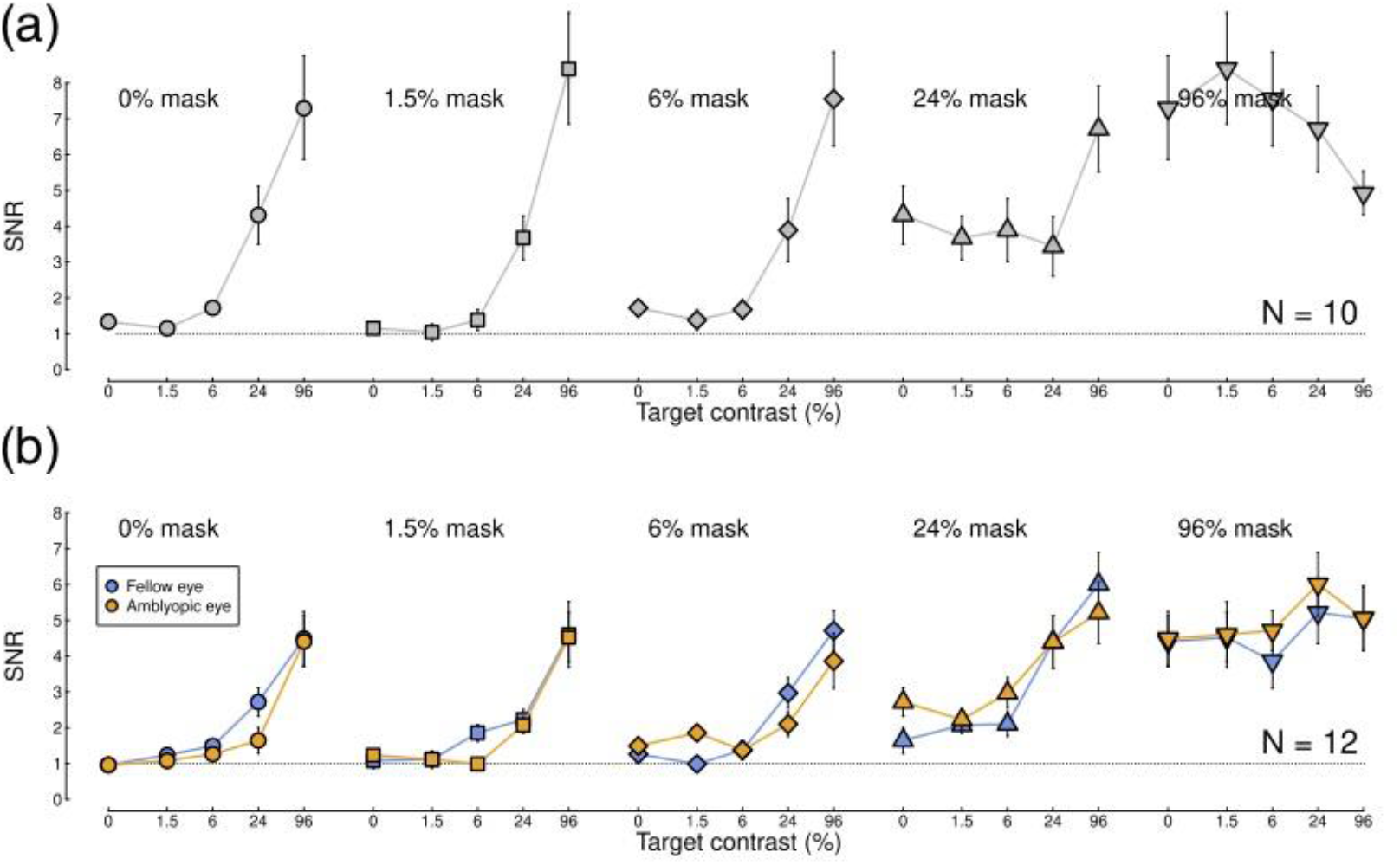
SSVEP data for only the participants who completed the fMRI experiment. The main trends are the same as in Figure 5c,d. Panel (a) shows data from control participants (N=10), and panel (b) shows data from patients (N=12).

